# Evaluating approaches of training a generative large language model for multi-label classification of unstructured electronic health records

**DOI:** 10.1101/2024.06.24.24309441

**Authors:** Dinithi Vithanage, Chao Deng, Lei Wang, Mengyang Yin, Mohammad Alkhalaf, Zhenyu Zhang, Yunshu Zhu, Alan Christy Soewargo, Ping Yu

## Abstract

Multi-label classification of unstructured electronic health records (EHR) is challenging due to the semantic complexity of textual data. Identifying the most effective machine learning method for EHR classification is useful in real-world clinical settings. Advances in natural language processing (NLP) using large language models (LLMs) offer promising solutions. Therefore, this experimental research aims to test the effects of zero-shot and few-shot learning prompting, with and without parameter-efficient fine-tuning (PEFT) and retrieval-augmented generation (RAG) of LLMs, on the multi-label classification of unstructured EHR data from residential aged care facilities (RACFs) in Australia. The four clinical tasks examined are agitation in dementia, depression in dementia, frailty index, and malnutrition risk factors, using the Llama 3.1-8B. Performance evaluation includes accuracy, macro-averaged precision, recall, and F1 score, supported by non-parametric statistical analyses. Results indicate that both zero-shot and few-shot learning, regardless of the use of PEFT and RAG, demonstrate equivalent performance across the clinical tasks when using the same prompting template. Few-shot learning consistently outperforms zero-shot learning when neither PEFT nor RAG is applied. Notably, PEFT significantly enhances model performance in both zero-shot and few-shot learning; however, RAG improves performance only in few-shot learning. After PEFT, the performance of zero-shot learning is equal to that of few-shot learning across clinical tasks. Additionally, few-shot learning with RAG surpasses zero-shot learning with RAG, while no significant difference exists between few-shot learning with RAG and zero-shot learning with PEFT. These findings offer crucial insights into LLMs for researchers, practitioners, and stakeholders utilizing LLMs in clinical document analysis.

## 1 Introduction

A substantial amount of medical predictive models have been trained, tested, and published, yet the majority of them have never been deployed into the clinical setting, which is coined “a last mile problem” [1]. This is because most of these predictive models are developed to handle structured health data, while much important clinical information is captured in free-text clinical notes, which introduces complexity for model development and deployment.

Electronic health records in RACFs in Australia are digitized systems designed to collect, store, and display data about clients’ demographics, medical diagnoses, assessments, progress notes, charts, and forms [1]. Similar to other healthcare settings [1], besides the structured diagnosis data, much important clinical information in RACFs is captured in unstructured, narrative, free-text nursing progress notes. Because free text is a more expressive and natural way for care staff to record care encounters and communicate among team members, these notes are often updated. They, thus, are the closest to real-time reflection of an older person’s health condition. Therefore, effectively extracting information from unstructured clinical notes in EHR is essential to support clinical decision-making, improve aged care quality, and advance translational research.

Multi-label classification assigns multiple labels or categories to a single input instance. Multi-label classification of free-text data is a specialized area in machine learning and natural language processing (NLP). It involves the automated extraction of entities, concepts, events, and their relations from unstructured text [2], a challenging task because text data often has different meanings and interpretations [3] and requires precise and expeditious information extraction [4]. Previous works in multi-label classification of EHR text using NLP have explored various approaches. Rule-based systems, including MERKI, MedLEE, SymText, NegEx, MedEx, MedXN, MetaMap, KnowledgeMap, HITEx, and ContextD, have been effectively utilized for this purpose [5–9]. These systems depend on predefined rules and specialized medical dictionaries to extract and categorize information from unstructured text. Although rule-based systems are known for their simplicity, their performance can decline if the narrative text includes terms not covered by the lexical resources [10].

Recently, algorithms like naïve Bayes, k-nearest neighbour, conditional random field, support vector machine, logistic regression, decision tree, random forest, and artificial neural networks have been employed for classification tasks in NLP [11]. Notably, transformer-based encoder-type language models, particularly various BERT models such as ClinicalBERT, SciBERT, and RoBERTa, have been applied to multi-label classification tasks using unstructured EHR data. For example, BERT models have been fine-tuned to handle the semantic complexity of EHR data, demonstrating effectiveness in assigning multiple labels to clinical notes [13]. Despite the advancement of various BERT models, Clinical NLP remains a labour-intensive process that demands a substantial amount of expertise and human efforts to prepare the training data [1, 12]. This limitation has hindered the practical application of the early NLP technique in information extraction from the unstructured, free-text EHR. Privacy and confidentiality concerns also hinder manual curation efforts and the sharing of annotated medical corpora [13].

The recent advancements in LLMs, such as GPT variants, T5, OPT, and Llama [2], have demonstrated the ability of these decoder models to generate text that is not only human-like but also surpasses human-level performance in specific tasks [2]. These models, when combined with machine learning techniques like pre-training, fine-tuning, retrieval augmented generation (RAG) and prompt-based learning [14], offer transformative potential for NLP, enabling the development of automated and adaptable systems that can extract valuable insights from the free-text EHR. This marks a significant step towards integrating health predictive models into real-world clinical systems.

We are still in the early days of applying generative AI-based LLMs to extract clinical insights from the free-text EHR. While LLMs have shown potential in answering clinical questions [4, 15, 16] and extracting clinical data from public health data sets [17], their practical application in specific tasks using clinical data produced in real-world clinical settings remains limited [4, 12, 17, 18]. It is yet to be determined whether prompt engineering for LLMs can meet the stringent safety standards required for healthcare applications, given their limitations in generating outputs that may contain disinformation, misinformation, bias or hallucinations [4, 19]. The optimal prompting strategies for healthcare information extraction, whether zero-shot or few-shot learning, using PEFT or RAG, in various contexts, remain unclear.

A prompt is an input a user enters to instruct an LLM to automatically generate sequential output [20]. An LLM uses pattern matching to identify the relationships between the words, phrases, and concepts in the prompt and connect these with the patterns learned from the previous training. It then uses natural language generation to respond in a human-understandable format. Prompts enable the model to adapt and comprehend specific information in a new domain, leveraging its learned knowledge stored within the pre-trained models like Llama 3, thereby expanding the model’s applicability and effectiveness. Prompt learning reduces the need to introduce new parameters or extensive retraining of the model using labelled data for various tasks, thus improving efficiency and reducing computational resources required for machine learning.

Extracting symptoms of various geriatric diseases is essential for symptom assessment, early disease diagnosis, personalised treatment, and improving patient outcomes. To date, there is no reporting of practical tools to execute this multi-label classification task accurately and reliably from free-text notes in an EHR system. There is also a lack of prior research on the difference in performance between zero-shot and few-shot learning for the same clinical classification task and the effect of PEFT or RAG on the tasks. As healthcare demands high safety standards for machine learning, it is imperative to conduct experimental comparisons of the performances of various machine learning methods. Understanding machine learning methods, e.g., prompting behaviour, is also crucial for the safe and effective deployment of LLMs in healthcare settings.

Therefore, we compared zero-shot and few-shot learning, with and without PEFT or RAG, on multi-label clinical classification tasks. In this study, we included four clinical tasks with careful consideration of the following factors: (1) the information is recorded in the free text nursing progress notes; (2) the information meets aged care information needs; and (3) the research team has curated labelled datasets to allow model training, validation, and testing to evaluate machine learning performance. We identified four clinical tasks: agitation in dementia, depression in dementia, frailty index and malnutrition risk factors (see Table 1). Each task has various numbers of labels, ranging from 13 to 83.

**Table 1:** Clinical tasks for multi-label classification.

| Clinical Tasks | Label Names | Number of Labels |
| --- | --- | --- |
| Agitation in dementia | Disruptive vocalisation, verbally aggressive behaviour, arguing, complaining, cursing, threat, using abusive language, using accusatory language, using foul language, using hostile language, using obscene language, using profane language, verbally nonaggressive behaviour, ceaseless talking, constant repetition of word, constant unwarranted requests for attention, constant unwarranted requests for help, constant unwarranted requests for reassurance, echolalia, groaning, grunting, howling, making bizarre noise, rambling, repetitive questioning, roaring, screaming, shouting, speaking in excessively loud voice, emotional distress, anger, frustration, irritability, mood swing, negativism, outburst, physically aggressive behaviour, biting, destroying property, fighting, grabbing, hitting, hurting self, hurting someone, kicking, pushing, resisting, scratching, shoving, slamming, spitting on people, staring, striking people, tearing, throwing object, physically nonaggressive behaviour, constant manipulation of object, fidgeting, gesturing, hand wringing, inappropriate dressing, inappropriate handling object, inappropriate undressing, pacing, pointing finger, repetitive physical mannerism, restlessness, rocking, rummaging, searching, wandering, bruxism, resisting, punching, absconding, | 83 |
|  | calling out, physical agitation, facial grimacing, moving furniture, hoard items, intrusive of others privacy, gets up and down from constantly, urinating on the floor |  |
| Depression in dementia | Diminished ability of thinking, feeling discouraged, feeling empty, feeling hopeless, feeling of excessive guilt, feeling sad, feeling worthless loss of energy, loss of interest, loss of pleasure, suicidal ideation, suicide, suicide attempt, tearfulness | 13 |
| Frailty index | Activity limitation, anaemia and haematinic deficiency, arthritis, atrial fibrillation, cerebrovascular disease, chronic kidney disease, diabetes, dizziness, dyspnoea, falls, foot problems, fragility fracture, hearing impairment, heart failure, heart valve disease, housebound, hypertension, hypotension/syncope, ischaemic heart disease, memory and cognitive impairment, mobility and transfer problems, osteoporosis, Parkinsonism and tremor, peptic ulcer, peripheral vascular disease, polypharmacy, requirement for care, respiratory disease, skin ulcer, sleep disturbance, social vulnerability, thyroid disease, urinary incontinence, urinary system disease, visual impairment, weight loss and anorexia | 36 |
| Risk factors for malnutrition | Anxiety, bowel blockage, cancer, chest infection, chronic wound, confusion, constipation, delirium, dementia, depression, diabetes, diarrhoea, difficulty swallow, dysphagia, eating disorder, food preference, frailty, gastritis, heart disease, HIV, hospital admission, isolation, kidney disease, liver disease, malabsorption medication, nausea, Parkinson, pneumonia, poor appetite, poor intake, poor oral health, pressure ulcer, sepsis, stroke, suboptimal intake, surgery, vomiting | 37 |

## 2 Methodology

We conducted the experiment in seven stages: generative AI-based large language model selection, data set selection, data preprocessing, designing prompt templates for zero-shot and few-short learning in each clinical task, machine learning methods execution, model performance evaluation and statistical analysis.

### 2.1 Ethics approval

The Human Research Ethics Committee of the University of Wollongong approved the study (Ethics Number 2019/159).

### 2.2 Generative AI-based LLM selection

We selected the Llama 3.1-8B-parameter model as the generative AI-based LLM. The selection considered the following factors: (1) the optimal model in terms of open source and favourable review at the time of the experiment; (2) practical considerations regarding the availability of GPU resources; (3) feasibility for local server deployment, convenience and control over usage; (4) compliance with health data privacy regulations in Australia; (4) the presence of diverse variants spawned through fine-tuning, including Alpaca, Baizem, Koala, and Vicuna [2]. We obtained the Llama 3.1 8B-parameter model from the Hugging Face repository (https://huggingface.co/meta-llama/Meta-Llama-3.1-8B-Instruct).

### 2.3 Data set selection

De-identified demographic data and free-text nursing progress notes were collected for the same population of older people living in 40 RACFs in New South Wales, Australia, from 2019 to 2021. The structured demographic information included masked sequence numbers for client de-identification, age, and gender. The unstructured nursing notes included nursing assessment and progress reporting. They documented clients’ daily activities, care staff’s clinical observations, assessments of client’s care needs (including risk factors), and carer interventions.

### 2.4 Data preprocessing

Text preprocessing involved the removal of URLs and non-textual characters, such as extra delimiters and empty spaces in the dataset. We made a choice not to exclude stop words because many of them, like “a,” “be,” “very,” “should,” etc., held semantic relevance to the content [21].

### 2.5 Designing prompt templates for zero-shot and few-short learning in each clinical task

First, we selected prompt-based training via zero-shot and few-shot learning.

#### 2.5.1 Zero-shot learning

Zero-shot learning uses single-prompt instruction to train LLMs for specific NLP tasks, directly applying previously trained models to predict both seen and unseen classes without using any labelled training instances [22]. Zero-shot learning has achieved impressive performance in a variety of NLP tasks, such as summarization, dialogue generation, and question-answering [4]. Ge et al. use zero-shot learning to extract six data elements from patients’ abdominal imaging reports using an API implementation of the OpenAI GPT-3.5 turbo LLM, achieving an overall high accuracy of 88.9% [23]. They find that the level of accuracy of zero-shot learning reduces with more complex use cases. Their findings prove the feasibility of using general-purpose LLMs to extract structured information from clinical data with minimal technical expertise.

#### 2.5.2 Few-shot learning

Few-shot learning, also coined as in-context learning, refers to the ability of LLMs to perform tasks guided by a small set of representative examples provided in the prompt [24, 25]. These in-context examples not only teach the LLM the mapping from inputs to outputs but also activate the LLM’s parametric knowledge. Only requiring a handful of labelled training examples is a clear advantage of few-shot learning, making it data-efficient and accessible to knowledge domain users without expertise in machine learning [24]. Few-shot learning is particularly useful in situations where annotating text data is not convenient or expensive. Domain experts can quickly create a generative AI system for a new task by only providing a few examples. Importantly, few-shot learning does not change the underlying model weights [26]. This allows for efficient adaptation to new tasks without risking the loss of previously learned knowledge. However, the performance of few-shot learning varies and is highly task-dependent [24]. Its accuracy is also sensitive to the choice of prompt templates and in-context examples. Prior research finds that using semantically similar in-context examples to those with prior success can significantly enhance the performance of few-shot learning [27].

We adopted the zero-shot and few-shot learning template developed by Abdallaha et al. [28] to construct our prompt (see Figure 1).

**Figure 1:**
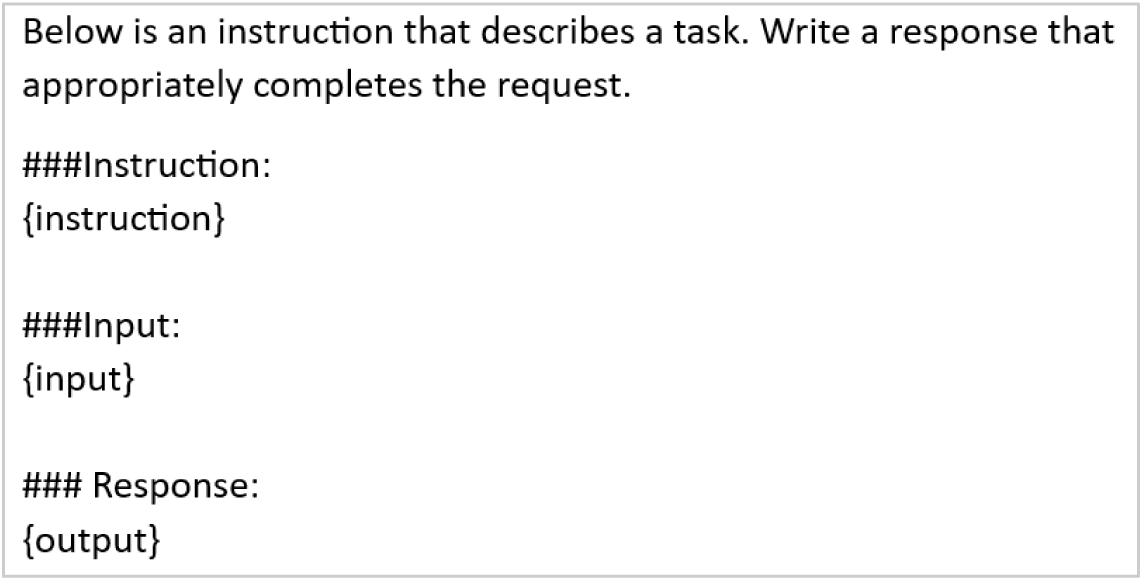
Prompt template adapted from Abdallaha et al. [28].

The final prompts used in our experiment are listed in Table 3. The example results generated from the final prompts are showcased in Supplementary Table 1.

**Table 2:** Research hypotheses in the study.

| Hypothesis | Null Hypothesis | Alternative Hypothesis |
| --- | --- | --- |
| 1 | Zero-shot and few-shot learning with similar prompting templates have the different level of performance when applied to multi-label classification for various clinical tasks. | Zero-shot and few-shot learning with similar prompting templates have the same level of performance when applied to multi-label classification for various clinical tasks. |
| 2 | Few-shot learning does not perform better than zero-shot learning for the multi-label classification of the same clinical task. | Few-shot learning performs better than zero-shot learning for the multi-label classification of the same clinical task. |
| 3 | Parameter-efficient fine-tuning does not improve the performance of either zero-shot or few-shot learning. | Parameter-efficient fine-tuning improves the performance of both zero-shot and few-shot learning. |
| 4 | Zero-shot learning does not reach the same level of performance as few-shot learning for the same clinical task after PEFT. | Zero-shot learning reaches the same level of performance as few-shot learning for the same clinical task after PEFT. |
| 5 | Fine-tuning for one clinical task does not impact model performance across other clinical tasks. | Fine-tuning for one clinical task impacts model performance across other clinical tasks. |
| 6 | Retrieval augmented generation does not improve the performance of both zero-shot and few-shot learning. | Retrieval augmented generation improves the performance of both zero-shot and few-shot learning. |
| 7 | Zero-shot learning does not reach the same level of performance as few-shot learning for the same clinical task with RAG. | Zero-shot learning reaches the same level of performance as few-shot learning for the same clinical task with RAG. |
| 8 | Few-shot learning with RAG and zero-shot learning with PEFT do not have the same performance levels in multi-label classification. | Few-shot learning with RAG and zero-shot learning with PEFT has the same performance levels in multi-label classification. |

**Table 3:** Prompts used in this study.

| Prompt Learning Technique | Domain | Prompt |
| --- | --- | --- |
| Zero-shot | Agitation in dementia | As a nursing expert, you are tasked with reviewing a nursing progress note for a resident with dementia residing in a Residential Aged Care environment. The note may contain one or more symptoms indicative of agitation in dementia, including but not limited to disruptive vocalisation, verbally aggressive behaviour, arguing, complaining, cursing, threat, using abusive language, using accusatory language, using foul language, using hostile language, using obscene language, using profane language, verbally nonaggressive behaviour, ceaseless talking, constant repetition of word, constant unwarranted requests for attention, constant unwarranted requests for help, constant unwarranted requests for reassurance, echolalia, groaning, grunting, howling, making bizarre noise, rambling, repetitive questioning, roaring, screaming, shouting, speaking in excessively loud voice, emotional distress, anger, frustration, irritability, mood swing, negativism, outburst, physically aggressive behaviour, biting, destroying property, fighting, grabbing, hitting, hurting self, hurting someone, kicking, pushing, resisting, scratching, shoving, slamming, spitting on people, staring, striking people, tearing, throwing object, physically nonaggressive behaviour, constant manipulation of object, fidgeting, gesturing, hand wringing, inappropriate dressing, inappropriate handling object, inappropriate undressing, pacing, pointing finger, repetitive physical mannerism, restlessness, rocking, rummaging, searching, wandering, bruxism, resisting, punching, absconding, calling out, physical agitation, facial grimacing, moving furniture, hoard items, intrusive of others privacy, gets up and down from constantly, urinating on the floor.<br>Please follow these steps.<br>1. Identify any symptoms of agitation.<br>2. If symptoms of agitation are evident, please list them. |
| Few-shot | Agitation in dementia | As a nursing expert, you are tasked with reviewing a nursing progress note for a resident with dementia residing in a Residential Aged Care environment. The note may contain one or more symptoms indicative of agitation in dementia, including but not limited to disruptive vocalisation, verbally aggressive behaviour, arguing, complaining, cursing, threat, using abusive language, using accusatory language, using foul language, using hostile language, using obscene language, using profane language, verbally nonaggressive behaviour, ceaseless talking, constant repetition of word, constant unwarranted requests for attention, constant unwarranted requests for help, constant unwarranted requests for reassurance, echolalia, groaning, grunting, howling, making bizarre noise, rambling, repetitive questioning, roaring, screaming, shouting, speaking in excessively loud voice, emotional distress, anger, frustration, irritability, mood swing, negativism, outburst, physically aggressive behaviour, biting, destroying property, fighting, grabbing, hitting, hurting self, hurting someone, kicking, pushing, resisting, scratching, shoving, slamming, spitting on people, staring, striking people, tearing, throwing object, physically nonaggressive behaviour, constant manipulation of object, |
|  |  | fidgeting, gesturing, hand wringing, inappropriate dressing, inappropriate handling object, inappropriate undressing, pacing, pointing finger, repetitive physical mannerism, restlessness, rocking, rummaging, searching, wandering, bruxism, resisting, punching, absconding, calling out, physical agitation, facial grimacing, moving furniture, hoard items, intrusive of others privacy, gets up and down from constantly, urinating on the floor.<br>Please follow these steps.<br>1. Identify any symptoms of agitation.<br>2. If symptoms of agitation are evident, please list them.<br>Example 1: You have identified that the resident exhibits agitation symptoms, including physical agitation/aggression and verbal behaviors, including verbal disruption, calling out, and screaming, as documented in the note below.<br>“Jenny, who suffers from vascular dementia, displays confusion, disorientation, and notable physical agitation/aggression and verbal behaviours. She exhibits verbal disruption, calling out and screaming, which disturbs others. Jenny frequently requires staff assistance for reassurance, comfort, and distraction using strategies such as music therapy, playing cards, or engaging in simple puzzles. Additionally, when highly distressed, staff contact her daughter to speak with her for comforting purposes. Although she can follow simple instructions, at times, she experiences considerable distress.”<br>Example 2: You have identified that the resident exhibits agitation symptoms, including wandering, frightening others, refusing care, and arguing, as documented in the note below.<br>"Elli faces challenges with poor balance and is at risk of falls due to impulsivity and reduced balance. Staff supervise Elli's transfers and mobility using a 4-wheeled walker. As Elli's dementia progresses, she tends to wander into other residents' rooms, mistakenly believing they occupy her bed. This behaviour frightens other residents, resulting in arguments. Staff frequently intervene, redirect Elli, and provide extra reassurance and diversion throughout the day. As her dementia advances, Elli lacks insight into her care needs. She adamantly refuses staff assistance changing her continence aids and attending to her hygiene. Due to this progression, staff guide her through mealtimes, set the table, provide cutlery, and supervise and encourage her during meals and drinks.” |
| Zero-shot | Frailty index | As a nursing expert, you are tasked with reviewing a nursing progress note for a resident residing in a Residential Aged Care environment. The note may contain one or more frailty index, including but not limited to activity limitation, anaemia and haematinic deficiency, arthritis, atrial fibrillation, cerebrovascular disease, chronic kidney disease, diabetes, dizziness, dyspnoea, falls, foot problems, fragility fracture, hearing impairment, heart failure, heart valve disease, housebound, hypertension, hypotension/syncope, ischaemic heart disease, memory and cognitive impairment, |
|  |  | mobility and transfer problems, osteoporosis, Parkinsonism and tremor, peptic ulcer, peripheral vascular disease, polypharmacy, requirement for care, respiratory disease, skin ulcer, sleep disturbance, social vulnerability, thyroid disease, urinary incontinence, urinary system disease, visual impairment, weight loss and anorexia.<br>Please follow these steps.<br>1. Identify any frailty index.<br>2. If the frailty index is evident, please list them along with the corresponding evidence in the note. |
| Few-shot | Frailty index | As a nursing expert, you are tasked with reviewing a nursing progress note for a resident residing in a Residential Aged Care environment. The note may contain one or more frailty index, including but not limited to activity limitation, anaemia and haematinic deficiency, arthritis, atrial fibrillation, cerebrovascular disease, chronic kidney disease, diabetes, dizziness, dyspnoea, falls, foot problems, fragility fracture, hearing impairment, heart failure, heart valve disease, housebound, hypertension, hypotension/syncope, ischaemic heart disease, memory and cognitive impairment, mobility and transfer problems, osteoporosis, Parkinsonism and tremor, peptic ulcer, peripheral vascular disease, polypharmacy, requirement for care, respiratory disease, skin ulcer, sleep disturbance, social vulnerability, thyroid disease, urinary incontinence, urinary system disease, visual impairment, weight loss and anorexia.<br>Please follow these steps.<br>1. Identify any frailty index.<br>2. If the frailty index is evident, please list them along with the corresponding evidence in the note.<br>Example 1: You have identified that the resident exhibits a frailty index, including mobility and transfer problems, based on the evidence that the resident mobilises with a wheelie walker, as documented in the note below.<br>"Ethan utilises a wheelie walker for mobility, and staff members provide supervision during his use of a chair lift. All of Ethan's meals take place in the dining room, with staff members responsible for pouring his drinks. Despite these assistance needs, Skeet maintains a healthy appetite."<br>Example 2: You have identified that the resident exhibits a frailty index, including a skin ulcer, based on the evidence that the resident's wound shows an unchanged state with a small amount of exudate in the same cavity, as documented in the note below.<br>" Wound location: Left foot. Date: 04/01/2020. Evaluation: The wound has remained unchanged since 02/01/2020, with the same cavity showing small exudate. The resident reports no pain except when the dressing is attended to. Scheduled for a specialist review next week." |
| Zero-shot | Depression in dementia | As a nursing expert, you are tasked with reviewing a nursing progress note for a resident with dementia residing in a Residential Aged Care environment. The note may contain one or more symptoms of depression, including but not limited to diminished ability of thinking, feeling discouraged, feeling empty, feeling hopeless, feeling of |
|  |  | excessive guilt, feeling sad, feeling worthless loss of energy, loss of interest, loss of pleasure, suicidal ideation, suicide, suicide attempt, tearfulness.<br>Please follow these steps.<br>1. Identify any symptoms of depression.<br>2. If symptoms of depression are evident, please list them. |
| Few-shot | Depression in dementia | As a nursing expert, you are tasked with reviewing a nursing progress note for a resident with dementia residing in a Residential Aged Care environment. The note may contain one or more symptoms of depression, including but not limited to diminished ability of thinking, feeling discouraged, feeling empty, feeling hopeless, feeling of excessive guilt, feeling sad, feeling worthless loss of energy, loss of interest, loss of pleasure, suicidal ideation, suicide, suicide attempt, tearfulness.<br>Please follow these steps.<br>1. Identify any symptoms of depression.<br>2. If symptoms of depression are evident, please list them.<br>Example 1: You have identified that the resident exhibits depression symptoms, including apathy and refusal of hygiene care, as documented in the note below.<br>"Due to Peter's depression, he is apathetic and refuses hygiene care. Staff need to reapproach Peter multiple times per day and spend extra time with him, providing encouragement and reassurance and explaining the importance of attending to hygiene care. He enjoys mass and music."<br>Example 2: You have identified that the resident exhibits a depression symptom, including a lack of motivation, as documented in the note below.<br>"Due to John's depression, John does not want to mobilise using mobility aids. He does not want to use a 4-wheeled walker or walking stick. Staff need to help get him up from bed and help sit him in a wheelchair for meals or outings after persuasion and encouragement." |
| Zero-shot | Malnutrition risk factors | As a nursing expert, you are tasked with reviewing a nursing progress note for a resident residing in a Residential Aged Care environment. The note may contain one or more malnutrition risk factors, including but not limited to anxiety, bowel blockage, cancer, chest infection, chronic wound, confusion, constipation, delirium, dementia, depression, diabetes, diarrhoea, difficulty swallow, dysphagia, eating disorder, food preference, frailty, gastritis, heart disease, HIV, hospital admission, isolation, kidney disease, liver disease, malabsorption medication, nausea, Parkinson, pneumonia, poor appetite, poor intake, poor oral health, pressure ulcer, sepsis, stroke, suboptimal intake, surgery, vomiting.<br>Please follow these steps.<br>1. Identify any risk factors of malnutrition.<br>2. If risk factors of malnutrition are evident, please list them. |
| Few-shot | Malnutrition risk factors | As a nursing expert, you are tasked with reviewing a nursing progress note for a resident residing in a Residential Aged Care environment. The note may contain one or more malnutrition risk factors, including but not limited to As a nursing expert, you are tasked with reviewing a nursing progress note for a resident residing in a Residential Aged Care environment. The note may contain one or more malnutrition risk factors, including but not limited to anxiety, bowel blockage, cancer, chest infection, chronic wound, confusion, constipation, delirium, dementia, depression, diabetes, diarrhoea, difficulty swallow, dysphagia, eating disorder, food preference, frailty, gastritis, heart disease, HIV, hospital admission, isolation, kidney disease, liver disease, malabsorption medication, nausea, Parkinson, pneumonia, poor appetite, poor intake, poor oral health, pressure ulcer, sepsis, stroke, suboptimal intake, surgery, vomiting.<br>Please follow these steps.<br>1. Identify any risk factors of malnutrition.<br>2. If risk factors of malnutrition are evident, please list them.<br>Example 1: You have identified that the resident exhibits a malnutrition risk factor, including confusion, as documented in the note below.<br>" John requires comprehensive assistance with his hygiene and toileting needs due to his confusion. He experiences incontinence of both urine and feces, necessitating the use of pads around the clock. A nurse is responsible for assisting him with toileting, changing his pads, cleansing his groin area, and applying barrier cream to mitigate the risk of skin issues or breakdown."<br>Example 2: You have identified that the resident exhibits a malnutrition risk factor, including constipation, as documented in the note below.<br>" Peter requires the assistance of a nurse for his hygiene and toileting needs, primarily due to his unsteady gait. He utilises pads due to incontinence issues. Additionally, he is prone to constipation and receives aperients as necessary. The current intervention measures in place have proven to be effective." |

### 2.6 Machine learning methods execution

We selected Llama 3.18B with PEFT, prompt-based learning (without PEFT or RAG), and RAG to test the LLM’s ability to adapt, generalize, and optimize performance in clinical multi-domain classification tasks.

#### 2.6.1 Experiment setup

##### Parameter efficient fine-tuning with LoRA on Llama 3

Parameter-efficient fine-tuning involves modifying the LLM, or the parameters used to train the LLM, to improve model response to the same prompt [20]. Fine-tuning changes a model’s weight, thus, the model’s behavior to perform better at a specific task. Full fine-tuning will fine-tune all layers of the pre-trained model, which can be computationally expensive and may lead to catastrophic forgetting, i.e., the model forgets the knowledge it gained during pre-training. Thus, it may significantly increase the cost of computational resources and computational skill sets. Parameter-efficient fine-tuning only fine-tunes a small number of (extra) parameters while freezing most parameters of the pre-trained LLMs. It thus overcomes the computational resource constraint and catastrophic forgetting observed in the full-scope fine-tuning of LLM.

Low-Rank Adaptation (LoRA) is a PEFT technique designed to improve training efficiency for LLMs. It freezes the weight of per-trained LLMs and inserts low-rank decomposition matrices into the transformer layers. Previous research has demonstrated that LoRA can allow the fine-tuning process to focus on crucial parameters specific to the target task or domain, thus optimizing the model’s performance without extensive resource requirements or overfitting concerns. By focusing on PEFT, LoRA minimizes the dependency on extensive labelled data for model optimization, which maximizes the utility of available data, making the fine-tuning process more effective and feasible in scenarios with limited annotated datasets [29].

We used the PEFT method to fine-tune the Llama model. The experiment was conducted on four NVIDIA RTX - A5000, each equipped with 24GB of memory. The use of multiple GPUs not only accelerated the training process but also ensured that the model could be fine-tuned within a reasonable timeframe. Our software environment was Ubuntu 18.04, the programming language was Python 3.10.0, and the deep learning framework was Pytorch 2.0.0. Instruction data points were employed during the PEFT process, and the hyperparameter settings are listed in Table 4. In these hyperparameter settings, batch size refers to the number of training examples processed in a single iteration of the model’s training process. However, due to the memory limitations of the available hardware, we employed a micro-batch size strategy, which splits the full batch into smaller chunks and processes them sequentially. This allowed us to handle the memory load efficiently while still achieving the target batch size of 128 with the Llama model [30, 31]. To further optimize memory usage, we applied 8-bit quantization using the BitsandBytes library, compressing the model weights into a lower precision format that fits within our GPU memory [32]. This same quantization strategy was utilized for other experiments as well, including (1) prompt-based learning without PEFT or RAG and (2) RAG, which are discussed in subsequent sections. The combination of micro-batch size and quantization was essential for effectively training the model within the hardware constraints, ensuring the smooth and efficient execution of our experiments.

**Table 4:** Hyperparameters used in PEFT.

| Settings | Parameters |
| --- | --- |
| Batch size | 128 |
| Micro batch size | 4 |
| LoRA rank | 8 |
| LoRA alpaca | 16 |
| LoRA dropout | 0.05 |
| Learning rate | 3e-4 |
| Training steps | 300 |
| Optimizer | AdamW |
| Trainable parameters (%) | <0.01% |

Llama 3 model’s maximum token limit is 8192, which was large enough to encompass the available tokens for each nursing note. During the fine-tuning process, the model iteratively processed each note within the defined token limit. The labelled dataset used in our study was originally annotated following the procedures detailed in Zhu et al. (2024) by the large research group [33]. The annotated data set for agitation in dementia was developed for the study of Zhu et al. (2023) [34], the malnutrition data set was developed for the study of Alkhalaf et al (2023) [35]. Applying the same manual annotation method to the same data set, we further developed annotation datasets for depression in dementia and frailty index. We randomly divided this labelled data (ensuring no overlapping free-text notes within the labelled dataset) detailed in Table 5 into 80% training, 10% validation, and 10% testing sets for each clinical task. This process was repeated three times to mitigate potential bias from different data splits, and cross-validation was conducted to achieve reliable results [36]. The dedicated test datasets were explicitly used to assess the other machine learning methods including prompt-based learning (without PEFT or RAG), and RAG, allowing for a comprehensive comparison and analysis of each clinical task to test the research hypotheses outlined in Table 2.

**Table 5:** Number of labelled data and file size for each clinical task.

| Clinical Tasks | Training + Validation Data | File Size | Output model |
| --- | --- | --- | --- |
| Agitation in dementia | 3000 nursing notes | 5.89 MB | Agitation in dementia with specialized PEFT of Lama 2 |
| Depression in dementia | 700 nursing notes | 280 KB | Depression in dementia with specialized PEFT of Lama 2 |
| Frailty index | 949 nursing notes | 154 KB | Frailty index with specialized PEFT of Lama 2 |
| Malnutrition risk factors | 2850 nursing notes | 972 KB | Malnutrition risk factors with specialized PEFT of Lama 2 |

We used the prompts delineated in Table 3 to evaluate the test data. First, we conducted zero-shot learning with or without PEFT or RAG on Llama 3 across the test datasets for each clinical task. This was followed by few-shot learning with or without PEFT or RAG on Llama 3 across the same test datasets for each clinical task. To ensure that few-shot learning does not benefit from any residual effects of the previous zero-shot learning during testing, we began each time with the downloaded Llama 3 model from the Hugging Face repository for each training method and clinical task. Furthermore, to address variability in results caused by model randomness, multiple evaluation sessions were conducted over three-weeks, with results being collected at three distinct time points. We applied the same approach of repeated evaluation sessions for consistency in two other experiments—prompt training without PEFT or RAG, and RAG—allowing for a more reliable and comparable evaluation across all methods.

##### Prompt-based learning with zero-shot and few-shot learning on Llama 3

The experiment was conducted in an environment similar to the one described in the “Parameter efficient fine-tuning with LoRA on Llama 3” section, with the main difference being the number of GPUs used. Since prompt-based learning with the original Llama 3 requires fewer computational resources due to the absence of fine-tuning [37], we were able to utilize a single GPU with 24GB. The same process was employed for the experiment with RAG, as no fine-tuning was involved in the RAG process. We employed Llama 3, utilizing zero-shot and few-shot learning prompts as outlined in Table 3. To prevented model contamination, we approached each clinical task (see Table 1) in two distinct steps. Initially, we employed the Llama 3 model that we directly downloaded from the Hugging Face repository, for zero-shot learning. Afterwards, we downloaded a new copy of the same model from the same repository for few-shot learning. The Llama 3 model has a maximum token limit of 8192, and none of the test notes (see the sections titled ‘parameter efficient fine-tuning with LoRA on Llama 3’ for details about the test datasets) exceeded this token count during evaluation. Consequently, the model processed each note iteratively within this defined token limit during testing.

##### Retrieval augmented generation with Llama 3

Retrieval-augmented generation enhances LLMs by enabling access to real-time, relevant information from external knowledge sources, improving response accuracy and relevance [32]. The process includes indexing, where data is converted into embeddings for efficient retrieval; retrieval, where the system identifies the most pertinent documents based on a user query; augmentation, where the retrieved information enriches the query’s context; and generation, where the LLM produces a response informed by both the original query and the augmented data. This dynamic approach addresses traditional LLM limitations, ensuring up-to-date and contextually accurate responses [32].

The experiment for Llama 3, with RAG, began with data preparation, utilizing a JSON file containing nursing notes that encompass all the labels for each clinical task (refer to Table 1). Each clinical task, and the number of nursing notes provided in the JSON file, are detailed in Table 6.

**Table 6:** Data used in the RAG.

| Clinical Tasks | Number of nursing notes | File Size |
| --- | --- | --- |
| Agitation in dementia | 83 | 14KB |
| Depression in dementia | 13 | 5 KB |
| Frailty index | 36 | 8 KB |
| Malnutrition risk factors | 37 | 10 KB |

This data was input into an embedding model. The embedding model (https://huggingface.co/sentence-transformers/all-mpnet-base-v2) generated embeddings from the data, which were then stored in a vector store. A query was formulated by combining the prompt with zero-shot or few-shot examples. The query was applied to the dedicated test datasets (see the sections titled ‘parameter efficient fine-tuning with LoRA on Llama 3’ for details about the test datasets). The retriever used the query to fetch relevant data from the vector store. The retrieval process, managed by LlamaIndex, determined relevance based on the similarity between the query and the stored embeddings, which were then used by the LLM to generate the output [38, 39].

### 2.7 Model performance evaluation

To assess model performance on the four clinical tasks, we utilized micro-average precision, recall, F1 score, and accuracy, which were more appropriate for imbalanced class distributions [40, 41]. For the automatic evaluation of multi-label text classification, we first calculated the BERTscore to obtain precision, recall, and F1 score following the method by Li et al. [42]. We then used the Scikit-learn library, which offers built-in metrics for multi-label classification, to calculate the micro-average F1 score, precision, and recall [43, 44]. The model outputs, generated in JSON format, were compared against the annotated ground truth data, also stored in JSON format. An extracted entity or phrase was considered correct if it overlapped the text and conveyed the exact or highly similar meaning of the annotated ground truth entity or phrase. The comparison was facilitated by BERTscore and Scikit-learn library tools. For instance, if the ground truth annotation is ‘shouting’ as a symptom of agitation in dementia, and the model output is also ‘shouting,’ it is evaluated as an exact match. Alternatively, if the ground truth annotation is ‘reject meals’ and the model output is ‘refuse meals,’ it is considered a match based on semantic similarity, as the two phrases share the same meaning. To measure semantic similarity between model predictions and the ground truth, a similarity threshold of 0.5 was used, following prior work in this domain [45, 46].

### 2.8 Statistical analysis

As the measurement indicators including accuracy, precision, recall and F1 score were not normally distributed, we utilized the non-parametric Kruskal-Wallis test for comparing results across three or more independent groups and the Mann-Whitney U test for comparing two independent groups to test the hypotheses, as suggested by the previous research [28, 47]. A significant difference is decided if the p-value is smaller than 0.05. The results generated from the statistical analysis are showcased in Supplementary Table 2.

## 3 Results

### 3.1 Results of testing Hypothesis 1: Zero-short and few-shot learning with similar prompting templates may (or may not) exhibit the same level of performance when applied to multi-label classification for various clinical tasks

To evaluate Hypothesis 1, we compared: (1) the performance of zero-shot learning (without PEFT or RAG); (2) the performance of zero-shot learning with PEFT; (3) the performance of zero-shot learning with RAG; (3) the performance of few-shot learning (without PEFT or RAG); (5) the performance of few-shot learning with PEFT; and (6) the performance of few-shot learning with RAG across the four clinical tasks.

#### 3.1.1 Comparing the performance of zero-shot learning (without PEFT or RAG) for four clinical tasks

There is no statistically significant difference in accuracy, precision, recall, and F1 score between these classification tasks undertaking this training method (Figure 2-a, p >0.05). However, there is a trend that the classification tasks related to agitation in dementia and malnutrition risk factors perform better than those related to frailty index and depression in dementia.

**Figure 2:**
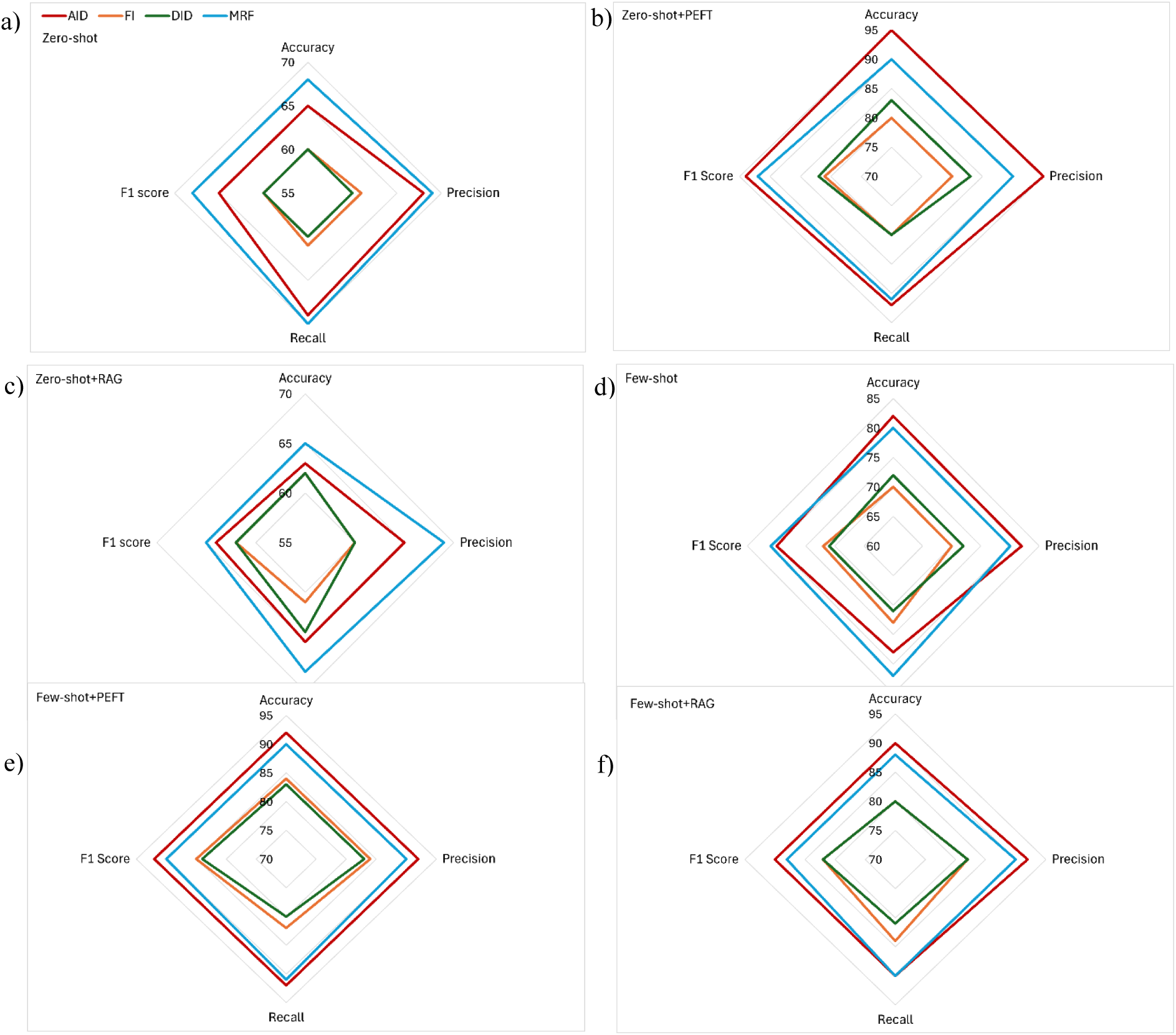
Comparative evaluation of model performance for four multi-label classification tasks with the following training methods: (a) zero-short learning, (b) zero-short learning with PEFT, (c) zero-short learning with RAG, (d) few-short learning, (d) few-short learning with PEFT, and (f) few-short learning with RAG. Note: ‘AID’ denotes agitation in dementia, ‘FI’ denotes frailty index, ‘DID’ denotes depression in dementia, ‘MRF’ denotes malnutrition risk factors. The same notation applies to the figures that follow. Note: ‘+PEFT’ denotes with PEFT. The same notation applies to the figures that follow. Note: ‘+RAG’ denotes with RAG. The same notation applies to the figures that follow.

#### 3.1.2 Comparing the performance of zero-shot learning with PEFT for four clinical tasks

Once again, no statistically significant difference is found in accuracy, precision, recall, and F1 score between these classification tasks undertaking this training method (Figure 2-b, p >0.05). However, there is a trend that the classification tasks related to agitation in dementia and malnutrition risk factors perform better than those related to frailty index and depression in dementia.

#### 3.1.3 Comparing the performance of zero-shot learning with RAG for four clinical tasks

No statistically significant difference is found in accuracy, precision, recall, and F1 score between these tasks undertaking this training method (Figure 2-c, p >0.05). However, the same trend as above is found, i.e., the classification tasks related to agitation in dementia and malnutrition risk factors perform better than those related to frailty index and depression in dementia.

#### 3.1.4 Comparing the performance of few-shot learning (without PEFT or RAG) for the four clinical tasks

No statistically significant difference is found in accuracy, precision, recall, and F1 score between these tasks undertaking this training method (Figure 2-d, p >0.05). However, the same trend as above is found, i.e., the classification tasks related to agitation in dementia and malnutrition risk factors perform better than those related to frailty index and depression in dementia.

#### 3.1.5 Comparing the performance of few-shot learning with PEFT for the four clinical tasks

Again, no statistically significant difference is found in accuracy, precision, recall, and F1 score among the four clinical tasks (Figure 2-e, p >0.05); however, the same trend as above is observed, i.e., the classification tasks related to agitation in dementia and malnutrition risk factors perform better than those related to frailty index and depression in dementia.

#### 3.1.6 Comparing the performance of few-shot learning with RAG for four clinical tasks

No statistically significant difference is found in accuracy, precision, recall, and F1 score between these tasks undertaking this training method (Figure 2-f, p >0.05). However, the same trend as above is found, i.e., the classification tasks related to agitation in dementia and malnutrition risk factors perform better than those related to frailty index and depression in dementia.

### 3.2 Results of testing Hypothesis 2: Few-shot learning may (or may not) perform better than zero-shot learning for the multi-label classification of the same clinical tasks

To evaluate Hypothesis 2, we compared the performance of zero-shot and few-shot learning (without PEFT and RAG) for all four clinical tasks. Few-shot learning significantly improves model accuracy, precision, recall and F1 score in all four multi-label clinical classification tasks than zero-shot learning (Figure 3-a, p < 0.05). The level of improvement includes an 18% increase in model accuracy, an 18% increase in precision, a 25% increase in recall, and a 28% increase in F1 score.

**Figure 3:**
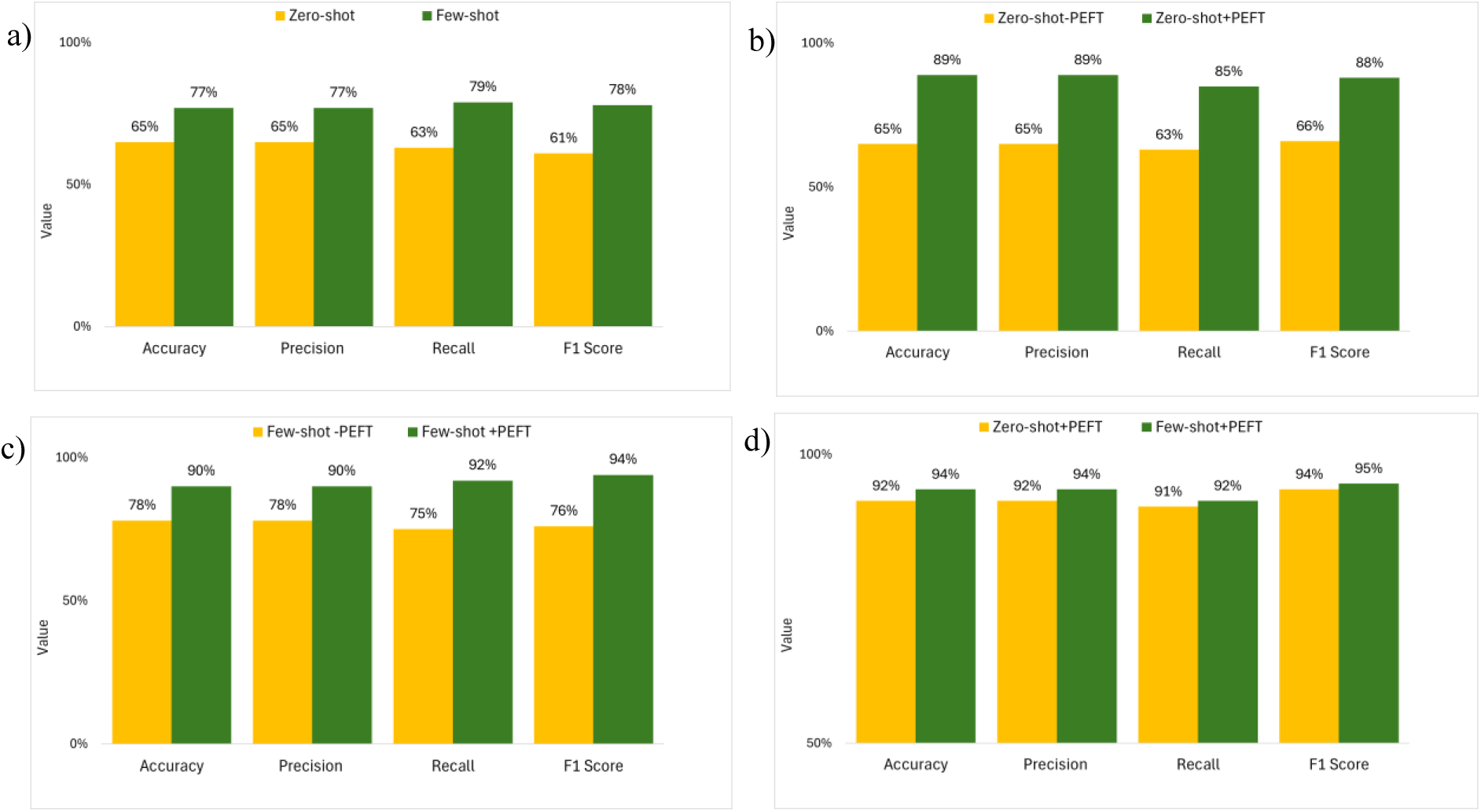
Comparative evaluation of model performance for four multi-label classification tasks with the following training methods: (a) zero-shot learning versus few-shot learning, (b) zero-shot learning versus zero-shot learning with PEFT, (c) few-shot learning versus few-shot learning with PEFT, and (d) zero-shot learning versus few-shot learning with PEFT. Note: ‘-PEFT’ denotes without PEFT. The same notation applies to the figures that follow.

### 3.3 Results of testing Hypothesis 3: Parameter-efficient finetuning may (or may not) improve both zero-shot and few-shot learning performance

To evaluate Hypothesis 3, we compared (1) the performance of the zero-shot learning model without PEFT and with PEFT; and (2) the performance of the few-shot learning model without PEFT and with PEFT for all four tasks.

#### 3.3.1 Comparing the performance of zero-shot learning model without PEFT and with PEFT for all four tasks

Zero-shot learning with PEFT significantly improves model accuracy, precision, recall and F1 score in all four multi-label clinical classification tasks (Figure 3-b, p < 0.05). The level of improvement is as follows: a 37% increase in model accuracy, a 37% increase in precision, a 35% increase in recall, and a 33% increase in F1 score.

#### 3.3.2 Comparing the performance of the few-shot learning model without PEFT and with PEFT for all four multi-label clinical classification tasks

Few-shot learning with PEFT significantly improves model accuracy, precision, recall and F1 score in all four multi-label clinical classification tasks (Figure 3-c, p < 0.05). The level of improvement is as follows: a 15% increase in model accuracy, a 15% increase in precision, a 23% increase in recall, and a 24% increase in F1 score.

### 3.4 Results of testing Hypothesis 4: Zero-shot learning may (or may not) reach the same level of performance as few-shot learning for the same clinical task with PEFT

To evaluate Hypothesis 4, we compared the performance of zero-shot and few-shot learning with PEFT for all four tasks. Although no statistically significant difference is found in accuracy, precision, recall, and F1 score between the zero-shot and few-shot learning with PEFT in all four multi-label clinical classification tasks (Figure 3-d, p >0.05), there is a trend that few-shot learning performs above zero-shot learning.

### 3.5 Results of testing Hypothesis 5: Fine-tuning for one clinical task may (or may not) impact model performance across other clinical tasks

To evaluate Hypothesis 5, we compared the performance of a clinical task-specific PEFT model with zero-shot learning, and its impact across other clinical tasks. For example, we conducted PEFT on the clinical task of agitation in dementia. We then compared the model’s impact on multi-label classification for depression in dementia, frailty index and malnutrition risk factors. We then compared the performance of a clinical task-specific PEFT model with few-shot learning and its impact across other clinical tasks, following the same pattern we did for zero-shot learning.

#### 3.5.1 Comparing the performance of a clinical task-specific PEFT model with zero-shot learning and its impact on other clinical tasks

No significant difference is found in accuracy, precision, recall, and F1 score for the other group of clinical tasks between two training models, pure zero-shot learning and zero-shot learning with PEFT for a specific clinical task. (see Figure 4-a, Figure 4-b, p >0.05).

**Figure 4.**
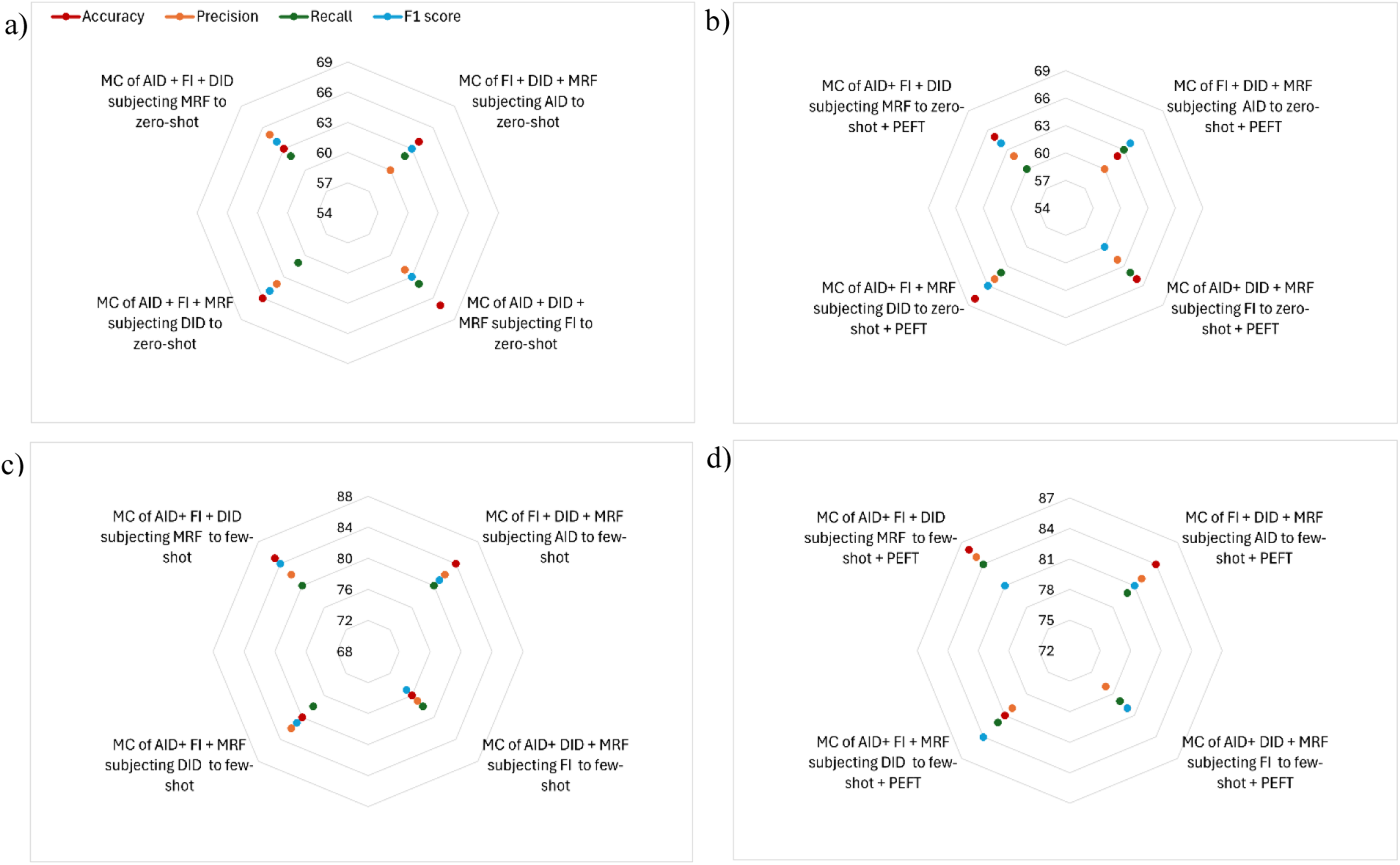
Performance of the measurement indicators for the other three multi-label classification tasks (a) when one clinical task was only trained with zero-shot learning, (b) when training on one clinical task with zero-shot learning and PEFT, (c) when one clinical task was only trained with few-shot learning, and (d) when training on one clinical task with few-shot learning and PEFT. Note: ‘MC’ denotes with multi-label classification.

#### 3.5.2 Comparing a clinical task-specific PEFT model performance with few-shot learning and its impact on other clinical tasks

No significant difference is found in accuracy, precision, recall, and F1 score for the other group of clinical tasks between two training models, pure few-shot learning and few-shot learning with PEFT for a specific clinical task. (see Figure 4-c, Figure 4-d, p >0.05).

### 3.6 Results of testing Hypothesis 6: Retrieval augmented generation may (or may not) improve both zero-shot and few-shot learning performance

To evaluate Hypothesis 6, we compared the performance of the model of (1) zero-shot learning without RAG and with RAG; and (2) few-shot learning without RAG and with RAG for all four tasks.

#### 3.6.1 Comparing the performance of zero-shot learning model without RAG and with RAG for all four tasks

Although no statistically significant difference is found in accuracy, precision, recall, and F1 score between the zero-shot learning without RAG and with RAG in all four multi-label clinical classification tasks (Figure 5-a, p >0.05), the results suggest a trend where zero-shot learning with RAG tends to perform better than without RAG.

**Figure 5.**
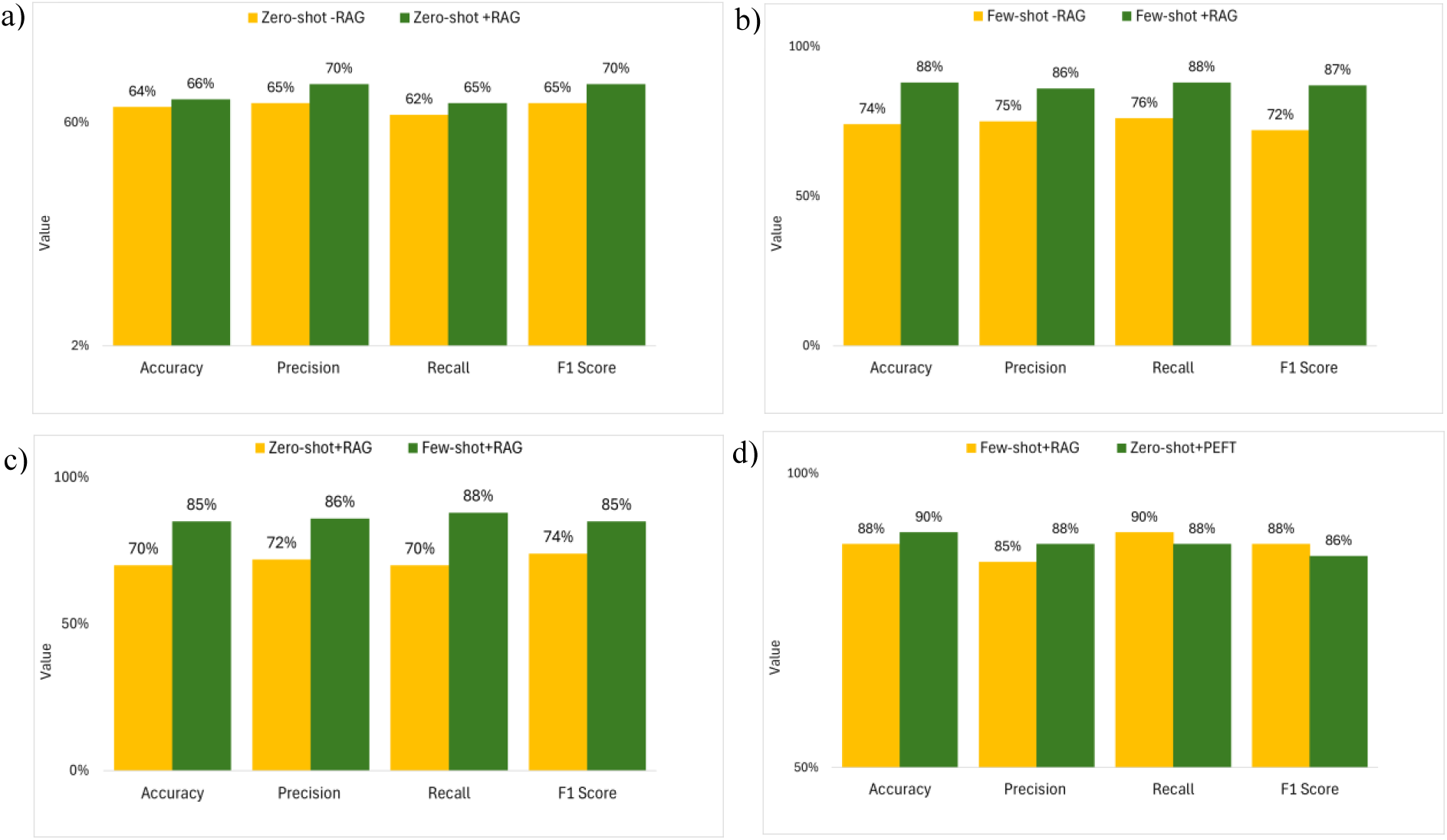
Comparative evaluation of model performance for four multi-label classification tasks with the following training methods: (a) zero-shot learning without RAG versus with RAG, (b) few-shot learning without RAG versus with RAG, (c) zero-shot learning with RAG versus few-shot learning with RAG, and (d) few-shot learning with RAG versus zero-shot learning with PEFT. Note: ‘-RAG’ denotes without RAG.

#### 3.6.2 Comparing the performance of few-shot learning model without RAG and with RAG for all four tasks

Few-shot learning with RAG significantly improves model accuracy, precision, recall and F1 score in all four multi-label clinical classification tasks (Figure 5-b, p < 0.05). The level of improvement is as follows: a 19% increase in model accuracy, a 15% increase in precision, a 16% increase in recall, and a 21% increase in F1 score.

### 3.7 Results of testing Hypothesis 7: Zero-shot learning may (or may not) reach the same level of performance as few-shot learning for the same clinical task with RAG

To evaluate Hypothesis 7, we compared the performance of zero-shot and few-shot learning with RAG for all four clinical tasks. Few-shot learning with RAG significantly improves model accuracy, precision, recall and F1 score in all four multi-label clinical classification tasks than zero-shot learning (Figure 5-c, p < 0.05). The level of improvement is as follows: a 17% increase in model F1 score, an 18% increase in model accuracy, an 18% increase in precision, and a 23% increase in recall.

### 3.8 Results of testing Hypothesis 8: Few-shot learning with RAG may (or may not) perform better than zero-shot learning with PEFT for all four tasks

To evaluate Hypothesis 8, we compared the performance of few-shot learning with RAG and zero-shot learning with PEFT for all four clinical tasks. No significant difference is found in accuracy, precision, recall, and F1 score between the few-shot learning with RAG and zero-shot learning with PEFT (Figure 5-d, p >0.05) in all four multi-label clinical classification tasks.

## 4 Discussion

This study explores the impact of zero-shot and few-shot prompt learning strategies, both with and without PEFT and RAG, on multi-label classification across four clinical tasks. These include agitation in dementia, depression in dementia, frailty index and malnutrition risk factors. To achieve this, eight research hypotheses have been formulated, and experimental designs have been implemented to rigorously test these hypotheses.

Our first hypothesis proposed that zero-shot and few-shot learning with similar prompting templates exhibit comparable performance in multi-label classification across various clinical tasks. Our findings support the alternative hypothesis, confirming that there is no significant difference in performance between these two learning approaches with similar prompting templates. While we did not identify statistically significant differences across all tasks, a consistent pattern emerged: the agitation in dementia and malnutrition risk factor classification tasks showed slightly better performance in accuracy, precision, recall, and F1 score compared to the frailty index and depression in dementia tasks. This performance disparity may be attributed to Llama 3 being trained on more data related to the former two tasks, leading to enhanced knowledge and better classification outcomes for those specific tasks.

Our second hypothesis proposed that few-shot learning performs better than zero-shot learning for the multi-label classification of the same clinical task. Our findings support the alternative hypothesis, confirming that few-shot learning significantly enhances performance compared to zero-shot learning (without PEFT or RAG). This improvement indicates that few-shot domain adaptation effectively minimizes false positives and false negatives while increasing true positives in classification tasks. These results highlight that exposing an LLM to initial information from the target domain can significantly improve its performance [48–50] in classification tasks. Our third hypothesis suggested that PEFT can improve both zero-shot and few-shot learning performance. The results support the alternative hypothesis, demonstrating that PEFT significantly improves performance in both learning modes compared to the models without PEFT. Based on our findings, we observed that applying PEFT within a specific domain effectively reduces false positives and false negatives while increasing true positives in domain-specific information extraction tasks. This confirms that the PEFT approach facilitates targeted modifications to the model’s parameters [27, 51, 52], leading to improved overall performance in multi-label classification tasks within that domain.

Our fourth hypothesis proposed that zero-shot learning can reach the same level of performance as few-shot learning for the same clinical task after PEFT. Our findings support the alternative hypothesis, confirming that there is no significant difference in performance between zero-shot and few-shot learning with PEFT. Based on our findings, this implies that further exposure through few-shot learning may not be necessary for models already fine-tuned within the domain. Our fifth hypothesis proposed that fine-tuning for one clinical task impacts model performance across other clinical tasks. However, the findings support the null hypothesis, indicating that fine-tuning for a specific task does not significantly affect the model’s performance on other classification tasks. The rationale behind this lies in the methodology of PEFT, which focuses on training only a selective subset of the pre-trained model’s parameters [27, 51]. This insight suggests that the model can be effectively tailored to a specific clinical task without compromising its effectiveness in handling diverse tasks. This adaptability underscores the potential of the PEFT approach within the LLM for various clinical tasks. Additionally, another factor for this result may be that the four clinical tasks are overall similar to each other in term of the nature.

Our sixth hypothesis asserts that RAG can improve both zero-shot and few-shot learning performance. However, the findings support the null hypothesis, showing no significant difference in performance between zero-shot learning with and without RAG. In contrast, a significant improvement was observed in few-shot learning when RAG was utilized. The lack of enhancement in zero-shot learning with RAG may be due to the absence of task-specific examples, which limits the model’s ability to effectively leverage retrieved information and generalize to unseen tasks [48, 49, 53]. Nevertheless, the results suggest that zero-shot learning with RAG consistently performs slightly better than without RAG, which could indicate that RAG offers a subtle advantage by providing additional context, though not enough to result in substantial gains. Conversely, the significant performance boost in few-shot learning suggests that RAG effectively leverages the limited examples provided to enrich model understanding, leading to improved classification accuracy. This disparity highlights RAG’s potential effectiveness in contexts where the model can benefit from supplementary information, particularly in few-shot settings [54, 55].

Our seventh hypothesis asserts that zero-shot learning can reach the same level of performance as few-shot learning for the same clinical task with RAG. Our findings support the null hypothesis, demonstrating that there is a significant difference in performance between zero-shot and few-shot learning with RAG. Zero-shot learning may not reach the same performance level as few-shot learning with RAG due to the absence of task-specific examples that help the model better understand the nuances of the clinical tasks. It relies solely on the model’s pre-existing knowledge [48, 49], which may not cover the specific clinical details required for accuracy. In contrast, few-shot learning provides the model with critical initial information and context from the target domain, enabling it to make more informed predictions [54–56]. As a result, while RAG can enhance contextual relevance, the lack of direct examples in zero-shot learning limits its effectiveness compared to few-shot approaches.

Our eighth hypothesis proposed that few-shot learning with RAG and zero-shot learning with PEFT achieve the same performance levels in multi-label classification. The results support the alternative hypothesis, showing that both methods can deliver similar outcomes despite their distinct mechanisms. Zero-shot learning with PEFT selectively fine-tunes the most relevant parameters [27], enabling the model to adapt effectively to new domains with minimal labelled data [27, 51]. On the other hand, few-shot learning with RAG benefits from integrating relevant external information through retrieval, combined with task-specific examples [55, 56]. This enhances the model’s ability to handle complex clinical data, leading to fewer classification errors, such as false positives and false negatives—an insight derived from our study’s findings. Our findings demonstrate that both PEFT and RAG excel in adapting to domain-specific tasks, highlighting their complementary strengths [57, 58] in improving model performance for multi-label classification in real-world clinical settings.

The findings of this study have significant implications for integrating AI models, particularly LLMs, into clinical practice. When combined both zero-shot and few-shot learning approaches with RAG and PEFT on multi-label classification tasks, the findings, present efficient solutions for improving multi-label classification in various healthcare settings. Given the ongoing challenge of obtaining labelled data from domain experts, RAG, combined with few-shot learning, emerges as an efficient option. By leveraging external knowledge sources through retrieval, RAG minimizes dependence on large, annotated datasets, reducing the need for extensive fine-tuning specific to each domain. This makes RAG a quicker and less resource-intensive solution, as it doesn’t require the same level of GPU resources or training time as PEFT, which do not require high technical expertise [37]. In contrast, PEFT demands more computational power and expertise, requiring substantial GPU resources and additional time for fine-tuning specific model parameters [37]. However, the enhanced performance that PEFT offers makes it useful in scenarios where the computational infrastructure is available.

This adaptability makes RAG equally valuable as PEFT for information retrieval in health and geriatric care, where clinical decision-making often relies on diverse and overlapping health conditions [59]. Therefore, this machine learning method can enable AI systems to integrate domain-specific knowledge in retrieving diagnostic and assessment data from the relevant but limited number of EHRs. This capability can improve diagnostic accuracy and support real-time health assessment, facilitating person-centred treatment and care plan development. Ultimately, the flexible deployment of this technique can help streamline care delivery in complex clinical contexts, reducing the cost and time associated with data labelling, requiring fewer resources and less training time, while maintaining robust model performance.

This study encompasses three notable limitations. Firstly, our study encompasses four multi-label clinical classification tasks. However, we recognize that these tasks may not represent a diverse spectrum of clinical scenarios. In the future, we will broaden our scope by incorporating additional clinical classification tasks into our study. Secondly, although we have examined four multi-label clinical classification tasks, additional tasks, such as question answering, summarization, and relation extraction are yet to be explored to enable a more comprehensive knowledge about the LLM’s performance in EHR data. The third limitation relates to the selection of model performance evaluation metrics, which utilizes accuracy, precision, recall, and F1 score as the primary evaluation metrics. In future studies, we will broaden our evaluation metrics to encompass calibration, robustness, fairness, bias, toxicity, and efficiency [60]. Diversifying evaluation criteria will provide a more comprehensive and nuanced assessment of the model’s performance across various dimensions. This will enhance our findings’ reliability and applicability in real-world EHR applications.

## 5 Conclusion

This study compares the performance of zero-shot and few-shot learning on multi-label clinical classification tasks and the impact of PEFT and RAG on their performance. Our findings reveal that the same prompting template—whether zero-shot or few-shot, with or without PEFT or RAG—yields comparable performance across different multi-label classification tasks. Few-shot learning consistently outperforms zero-shot learning in classification tasks, even without PEFT or RAG. However, when PEFT is integrated with both zero-shot and few-shot learning, zero-shot learning achieves performance levels similar to few-shot learning in classification tasks. On the other hand, with RAG, few-shot learning consistently surpasses zero-shot learning, indicating RAG’s superior ability to provide relevant external knowledge in few-shot learning. Our analysis also shows that fine-tuning LLMs for a particular clinical task does not significantly impact on the model’s performance when applied to other clinical tasks. Notably, few-shot learning with RAG achieves performance comparable to zero-shot learning with PEFT, further highlighting the adaptability and efficiency of these advanced methods in clinical applications.

## Data Availability

Data is not available.

## Acknowledgements

This research is part of a PhD project supported by the University of Wollongong, Australia and the University Grants Commission, Sri Lanka. It is also supported by Round 6 Research Grant from Aged Care Research and Innovation Grant Australia (ARIIA).

## Author Contributions

Conception of idea: [Dinithi Vithanage], [Ping Yu]. Literature search and data analysis: [Dinithi Vithanage], [Ping Yu]. Critical review and revision: [Dinithi Vithanage], [Ping Yu], [Lei Wang], [Chao Deng]. Data acquisition: [Mengyang Yin], Data annotation: [Mengyang Yin], [Mohammad Alkhalaf], [Zhenyu Zhang], [Yunshu Zhu], [Alan Christy Soewargo].

## Declarations

## Availability of Data and Material

Data is not available to comply with the Australian laws in privacy protection.

## Consent for Publication

All authors have approved the manuscript and agree with its submission to the Journal of Healthcare Informatics Research.

## Supplementary Material

The Supplementary Material for this article can be found in Supplementary Material.pdf.

## Statements and Declarations

This manuscript has not been published elsewhere and is not under consideration by another journal. All authors have approved the manuscript and agree with its submission to the Journal of Healthcare Informatics Research.

