## Supplemental Table for "Evaluating approaches of training a generative large language model for multi-label classification of unstructured electronic health records"

**Supplementary Table 1**

Example results generated from the final prompts.

| **Prompt** | **Input unstructured EHR text** | **Results from the model** |
| --- | --- | --- |
| As a nursing expert, you are tasked with reviewing a nursing progress note for a resident with dementia residing in a Residential Aged Care environment. The note may contain one or more symptoms indicative of agitation in dementia, including but not limited to disruptive vocalisation, verbally aggressive behaviour, arguing, complaining, cursing, threat, using abusive language, using accusatory language, using foul language, using hostile language, using obscene language, using profane language, verbally nonaggressive behaviour, ceaseless talking, constant repetition of word, constant unwarranted requests for attention, constant unwarranted requests for help, constant unwarranted requests for reassurance, echolalia, groaning, grunting, howling, making bizarre noise, rambling, repetitive questioning, roaring, screaming, shouting, speaking in excessively loud voice, emotional distress, anger, frustration, irritability, mood swing, negativism, outburst, physically aggressive behaviour, biting, destroying property, fighting, grabbing, hitting, hurting self, hurting someone, kicking, pushing, resisting, scratching, shoving, slamming, spitting on people, staring, striking people, tearing, throwing object, physically nonaggressive behaviour, constant manipulation of object, fidgeting, gesturing, hand wringing, inappropriate dressing, inappropriate handling object, inappropriate undressing, pacing, pointing finger, repetitive physical mannerism, restlessness, rocking, rummaging, searching, wandering, bruxism, resisting, punching, absconding, calling out, physical agitation, facial grimacing, moving furniture, hoard items, intrusive of others privacy, gets up and down from constantly, urinating on the floor.  Please follow these steps.   1. Identify any symptoms of agitation. 2. If symptoms of agitation are evident, please list them. | Maya experiences cognitive impairment due to dementia, resulting in a diminished ability to perform activities of daily living (ADLs) independently. Assistance is required for personal hygiene, toileting, and managing incontinence of both feces and urine. During mealtimes, Maya needs assistance and encouragement to eat, with occasional feeding as necessary. She adheres to a soft, bite-sized medium diet with thin fluids. Maya frequently engages in wandering behavior, necessitating gentle redirection. Additionally, she exhibits bruxism, often producing musical and repetitive sounds. | Based on the provided progress note, I have identified the following agitation symptoms:  wandering  bruxism  making musical, repeated sound |
| As a nursing expert, you are tasked with reviewing a nursing progress note for a resident residing in a Residential Aged Care environment. The note may contain one or more symptoms indicative of agitation in dementia, including but not limited to disruptive vocalisation, verbally aggressive behaviour, arguing, complaining, cursing, threat, using abusive language, using accusatory language, using foul language, using hostile language, using obscene language, using profane language, verbally nonaggressive behaviour, ceaseless talking, constant repetition of word, constant unwarranted requests for attention, constant unwarranted requests for help, constant unwarranted requests for reassurance, echolalia, groaning, grunting, howling, making bizarre noise, rambling, repetitive questioning, roaring, screaming, shouting, speaking in excessively loud voice, emotional distress, anger, frustration, irritability, mood swing, negativism, outburst, physically aggressive behaviour, biting, destroying property, fighting, grabbing, hitting, hurting self, hurting someone, kicking, pushing, resisting, scratching, shoving, slamming, spitting on people, staring, striking people, tearing, throwing object, physically nonaggressive behaviour, constant manipulation of object, fidgeting, gesturing, hand wringing, inappropriate dressing, inappropriate handling object, inappropriate undressing, pacing, pointing finger, repetitive physical mannerism, restlessness, rocking, rummaging, searching, wandering, bruxism, resisting, punching, absconding, calling out, physical agitation, facial grimacing, moving furniture, hoard items, intrusive of others privacy, gets up and down from constantly, urinating on the floor.  Please follow these steps.   1. Identify any frailty index. 2. If the frailty index is evident, please list them along with the corresponding evidence in the note.   Example 1: You have identified that the resident exhibits a frailty index, including mobility and transfer problems, based on the evidence that the resident mobilises with a wheelie walker, as documented in the note below.  "Ethan utilises a wheelie walker for mobility, and staff members provide supervision during his use of a chair lift. All of Ethan's meals take place in the dining room, with staff members responsible for pouring his drinks. Despite these assistance needs, Skeet maintains a healthy appetite."  Example 2: You have identified that the resident exhibits a frailty index, including a skin ulcer, based on the evidence that the resident's wound shows an unchanged state with a small amount of exudate in the same cavity, as documented in the note below.  " Wound location: Left foot. Date: 04/01/2020. Evaluation: The wound has remained unchanged since 02/01/2020, with the same cavity showing small exudate. The resident reports no pain except when the dressing is attended to. Scheduled for a specialist review next week." | In the 4-monthly physiotherapy care plan review conducted today, a mobility assessment was performed on Peter. The assessment indicates that Peter remains at his baseline, with no observed changes in his mobility requirements. Peter continues to necessitate supervision for bed mobility, transfers, and mobility when using a four-wheel walker (4WW). | Based on the review instructions, I have identified the following frailty symptoms:  Mobility and transfer problems, based on the evidence that Peter requires supervision for bed mobility and transfers, and mobility when using a four-wheel walker (4WW). |
| As a nursing expert, you are tasked with reviewing a nursing progress note for a resident with dementia residing in a Residential Aged Care environment. The note may contain one or more symptoms of depression, including but not limited to diminished ability of thinking, feeling discouraged, feeling empty, feeling hopeless, feeling of excessive guilt, feeling sad, feeling worthless loss of energy, loss of interest, loss of pleasure, suicidal ideation, suicide, suicide attempt, tearfulness.  Please follow these steps.   1. Identify any symptoms of depression. 2. If symptoms of depression are evident, please list them. | Asher, experiencing depression, expresses a reluctance to utilize mobility aids, such as a four-wheel walker (4WW) or walking stick. Consequently, staff members are required to provide physical assistance in mobilizing Asher, aiding him in getting up from bed and seating him in a wheelchair for meals or outings, following persuasion and encouragement. Leslie tolerated meals and drinks well today, and he is sitting in a lounge ATOR. | Based on the information provided, it appears that the resident is experiencing depression symptoms:  Lack of motivation or interest in activities, as indicated by Asher's reluctance to mobilize using mobility aids, such as a four-wheel walker (4WW) or walking stick. |
| As a nursing expert, you are tasked with reviewing a nursing progress note for a resident residing in a Residential Aged Care environment. The note may contain one or more malnutrition risk factors, including but not limited to anxiety, bowel blockage, cancer, chest infection, chronic wound, confusion, constipation, delirium, dementia, depression, diabetes, diarrhoea, difficulty swallow, dysphagia, eating disorder, food preference, frailty, gastritis, heart disease, HIV, hospital admission, isolation, kidney disease, liver disease, malabsorption medication, nausea, Parkinson, pneumonia, poor appetite, poor intake, poor oral health, pressure ulcer, sepsis, stroke, suboptimal intake, surgery, vomiting.  Please follow these steps.   1. Identify any risk factors of malnutrition. 2. If risk factors of malnutrition are evident, please list them.   Example 1: You have identified that the resident exhibits a malnutrition risk factor, including confusion, as documented in the note below.  " John requires comprehensive assistance with his hygiene and toileting needs due to his confusion. He experiences incontinence of both urine and feces, necessitating the use of pads around the clock. A nurse is responsible for assisting him with toileting, changing his pads, cleansing his groin area, and applying barrier cream to mitigate the risk of skin issues or breakdown.”  Example 2: You have identified that the resident exhibits a malnutrition risk factor, including constipation, as documented in the note below.  " Peter requires the assistance of a nurse for his hygiene and toileting needs, primarily due to his unsteady gait. He utilises pads due to incontinence issues. Additionally, he is prone to constipation and receives aperients as necessary. The current intervention measures in place have proven to be effective." | Kiya mobilizes with the assistance of a four-wheel walker (4WW) under staff supervision. Transfers are conducted with the supervision of one staff member to ensure safety, as Kiya is considered a fall risk due to her tendency to rush. Personal hygiene is assisted by one staff member due to confusion. | The nursing progress note indicates one risk factor of malnutrition for resident Kiya:  Confusion: The note mentions that Kiya requires assistance with personal hygiene due to confusion. |

**Supplementary Table 2**

Results generated from the statistical analysis.

| Method | Accuracy | Precision | Recall | F1 score |
| --- | --- | --- | --- | --- |
| Comparing the performance of zero-shot learning without PEFT or RAG for four clinical tasks | 0.36 | 0.31 | 0.34 | 0.33 |
| Comparing the performance of zero-shot learning with PEFT for four clinical tasks. | 0.08 | 0.09 | 0.08 | 0.075 |
| Comparing the performance of zero-shot learning with RAG for four clinical tasks. | 0.38 | 0.29 | 0.30 | 0.28 |
| Comparing the performance of few-shot learning without PEFT and RAG for the four clinical tasks. | 0.21 | 0.36 | 0.30 | 0.28 |
| Comparing the performance of few-shot learning with PEFT for the four clinical tasks. | 0.36 | 0.30 | 0.28 | 0.33 |
| Comparing the performance of few-shot learning with RAG for the four clinical tasks. | 0.38 | 0.32 | 0.36 | 0.39 |
| Comparing the performance of zero-shot and few-shot learning without PEFT and RAG for all four clinical tasks. | 3.2-7e | 2.8-5e | 2.0-7e | 1.5-7e |
| Comparing the performance of the model of zero-shot learning without PEFT and with PEFT for all four tasks. | 3.35-5e | 3.35-5e | 2.12 e-5e | 2.0-5e |
| Comparing the performance of the model of few-shot learning without PEFT and with PEFT for all four tasks. | 2.35-5e | 2.35-5e | 1.82-5e | 2.10-5e |
| Comparing the performance of zero-shot and few-shot learning with PEFT for all four tasks. | 2.32 | 2.32 | 1.85 | 1.84 |
| Comparing the performance of a clinical task-specialized PEFT model with zero-shot learning and its impact on other clinical tasks trained via zero-shot learning without PEFT. | 1.22 | 1.22 | 1.59 | 1.88 |
| Comparing the performance of a clinical task-specialized PEFT model with few-shot learning and its impact on other clinical tasks trained via few-shot learning without PEFT. | 0.8 | 0.8 | 1.22 | 0.99 |
| Comparing the performance of the model of zero-shot learning without RAG and with RAG for all four tasks. | 0.46 | 0.42 | 0.48 | 0.40 |
| Comparing the performance of the model of few-shot learning without RAG and with RAG for all four tasks. | 2.00-3e | 1.79-4e | 2.25-3e | 2.13-3e |
| Comparing the performance of zero-shot learning with RAG and few-shot learning with RAG. | 2.24-3e | 2.24-3e | 1.02-5e | 1.45-2e |
| Comparing the performance of few-shot learning with RAG and zero-shot learning with PEFT | 1.22 | 1.22 | 0.89 | 1.14 |
